# Impact of intra-partum Azithromycin on carriage of group A streptococcus in The Gambia: a posthoc analysis of a double-blind randomized placebo-controlled trial

**DOI:** 10.1101/2020.12.04.20236653

**Authors:** Isatou Jagne, Alexander J. Keeley, Abdoulie Bojang, Bully Camara, Edrissa Jallow, Elina Senghore, Claire Oluwalana, Saikou Y. Bah, Claire E. Turner, Abdul Karim Sesay, Umberto D’Alessandro, Christian Bottomley, Thushan I. de Silva, Anna Roca

**Affiliations:** Medical Research Council Unit the Gambia at the London School of Hygiene and Tropical Medicine, Banjul, The Gambia; Faculty of Infectious and Tropical Diseases, London School of Hygiene & Tropical Medicine, London, UK; The Florey Institute, University of Sheffield, Sheffield, UK

**Keywords:** Sub-Saharan Africa, azithromycin, bacterial carriage, group A *streptococcus*, *Streptococcus dysgalactiae subspecies equisimilis*

## Abstract

**Background:** Group A *Streptococcus* (GAS) is a major human pathogen and an important cause of maternal and neonatal sepsis.

**Methods:** We performed a posthoc analysis of a double-blind, placebo-controlled randomized-trial (ratio 1:1) carried out in The Gambia to determine the impact of one oral dose (2g) of intra-partum azithromycin on maternal and neonatal GAS carriage. Breast milk, nasopharyngeal and vaginal swabs were collected at different time points during 4 weeks post-treatment. All samples were processed using conventional microbiology techniques. Whole genome sequencing (WGS) of GAS isolates was performed by Illumina MiSeq platform.

**Results:** We randomized 829 mothers who delivered 843 babies. GAS carriage in mothers in the azithromycin arm was lower in breast milk (0.28% vs 2.48%, Prevalence Ratio (PR)=0.11, 95% CI 0.01-0.90) and the nasopharynx (0.28% vs 1.93%, PR=0.15, 95% CI 0.02-1.19), but not in the vaginal tract (1.99% vs 1.93%, PR=1.03, 95% CI 0.37-2.91). Among neonates, GAS carriage in the nasopharynx was slightly lower in the azithromycin arm (0.57% vs 1.91%, PR=0.30, 95% CI 0.06-1.42). Prevalence of azithromycin-resistant GAS was similar in both arms, except for a higher prevalence in the vaginal tract among women in the azithromycin arm (1.99% vs 0.28%, PR=7.24, 95% CI 0.87-56.92). WGS revealed ten of the 45 GAS isolates (22.2%) were *Streptococcus dysgalactiae* subspecies *equisimilis* expressing Lancefield group A carbohydrate (SDSE(A)). All SDSE(A) isolates were azithromycin-resistant, harbouring macrolide resistant genes *msrD* and *mefA*.

**Conclusions:** Oral intra-partum azithromycin reduced prevalence of GAS carriage among mothers and neonates. Azithromycin-resistant SDSE(A) carriage was observed among participants treated with azithromycin.

**Short Summary:** Group A streptococcus (GAS) is an important cause of sepsis. One oral dose (2g) of intra-partum azithromycin reduced maternal and neonatal GAS carriage. However, azithromycin-resistant *Streptococcus dysgalactiae* subspecies *equisimilis* expressing Lancefield group A carbohydrate was detected in women receiving azithromycin.

## BACKGROUND

Pregnant women and neonates are at highest risk of sepsis, up to 6 weeks after delivery, resulting in 6-19% of maternal deaths [1, 2]. Low- and middle-income countries (LMICs) have the highest burden of mortality due to maternal sepsis, with the highest rates in sub-Saharan Africa (SSA). Neonatal sepsis is a significant cause of severe morbidity and is estimated to cause 7% of under-5 mortality [3, 4]. Globally, *Staphylococcus aureus* and Group B *Streptococcus* (GBS) are main causes of maternal and neonatal sepsis [5-7]. Group A *Streptococcus* (GAS; *Streptococcus pyogenes*) is increasingly recognized as an important Gram-positive pathogen associated with maternal and neonatal sepsis [8-10]. GAS can cause both early and late onset of neonatal sepsis [11, 12], usually as a result of infection acquired through the birth canal [12].

Surveillance conducted in the USA between 1995 and 2000 estimated an annual incidence of 6 per 100,000 live births of GAS-related maternal sepsis, with a 3.5% case-fatality ratio in invasive disease [13]. A review of pregnancy-related GAS infections between 1974 and 2009 included no data from SSA [14]. Between 2010 and 2016, the incidence of GAS neonatal sepsis in London and the south of England was 1.5 per 100,000 person years [15]. There is limited data on the burden of GAS infections in SSA due to the lack of systematic surveillance [16]. In the Eastern Cape, South Africa, the mean annual incidence rate of invasive GAS infection was 6 cases per 100,000-person years in all age groups (58% of samples from 18-64 year olds) [17]. In Kenya, the incidence of neonatal GAS sepsis was 0.6 cases per 1,000 live births [18]. GAS also causes non-invasive disease, including tonsillo-pharyngitis, skin infections and rheumatic fever that can result in rheumatic heart disease [17, 19, 20].

We conducted a double-blind randomized trial (the PregnAnZI trial) to assess the effect of 2g of oral intra-partum azithromycin in reducing bacterial carriage of the main Gram-positive pathogens associated with neonatal sepsis (*S. aureus, Streptococcus pneumoniae* and GBS) in the mother and the newborn during the 4 weeks following the intervention [21]. The trial showed a consistent reduction of bacterial carriage of these three bacteria in both the mother and their newborns during the entire neonatal period and a decrease in the occurrence of disease [22, 23]. We present here a posthoc analysis of the trial to determine the effect of 2g intra-partum azithromycin on the prevalence and antibiotic resistance of GAS in mothers and their newborns during the 4 weeks following the intervention. We also used whole genome sequencing (WGS) to further characterise the GAS isolates and perform phylogenetic analysis.

## METHODS

### Study Design/Population

This study is a posthoc analysis of data from a double-blind, placebo-controlled randomized trial in which women in labour were randomized to receive either a single dose of 2g of oral azithromycin or placebo (ratio 1:1) [21]. The trial was conducted at the Jammeh Foundation for Peace (JFP), a government-run health facility located in western Gambia that manages approximately 4,500 deliveries per year. The population in the catchment area is representative of The Gambia and covers its main ethnic groups [22]. Between April 2013 and April 2014, women aged 18-45 years attending the JFP labour ward and with no acute or chronic conditions were recruited into the trial. Details of exclusion criteria have been reported elsewhere [21]. The women had provided written informed consent to participate in the study during previous antenatal care visits. Women and their newborns were followed for up to 8 weeks postpartum and biological samples were collected during the first 4 weeks [21, 22].

### Study Samples

A nasopharyngeal swab (NPS) and a low vaginal swab (VS) were collected from women before the intervention was administered and during labour. Post-intervention samples included: (i) newborn NPS within 6 h after birth; (ii) samples collected during home visits at days 3, 6, 14 and 28 (NPS from mothers and newborns, and breast milk(BM) from mothers) and (iii) a VS collected in the health facility during the postnatal check at day 8-10 post-delivery [22].

### Sample collection

NPS were collected by passing the tip of a calcium alginate (Expotech USA Inc) swab across the mucosa of the posterior wall of the nasopharynx. The swab was rotated and left in the nasopharynx for approximately 5 seconds. The inoculated swab was placed immediately into a vial containing skim milk-tryptone-glucose-glycerol (STGG) transport medium and then into a cold box before being taken to the Medical Research Council Unit The Gambia (MRC) at the London School of Hygiene and Tropical Medicine (LSHTM) laboratories within 8 hours of collection [21].

VS were collected by inserting a sterile cotton swab (Sterilin Ltd, UK) 2–3 cm into the vagina and rotating the swab with a circular motion, leaving it in the vagina for approximately 5 seconds. The inoculated swabs were then placed immediately into the vials containing STGG and put in a cold box before being transferred to the MRC laboratories within 8 hours [21].

Breast milk samples were collected by first disinfecting the nipple and areola of the breast using sterile cotton soaked with 0.02 % chlorhexidine. Mothers were then asked to manually express their milk. The first 0.5 mL was discarded. The following 1-2 mL was collected in a sterile plastic bijoux bottle put in a cold box and transferred to the MRC laboratories within 8 hours [21].

### Laboratory Procedures

#### GAS culture from NPS, VS and breast milk samples

Samples were vortexed for 20 seconds prior to storage at -70°C for subsequent processing in batches. During processing, samples were allowed to thaw on ice. Each vial was then vortexed briefly in order to homogenise the medium and 50μl was dispensed onto crystal violet blood agar (CVBA) (CM0085 Oxoid, UK +0.02 % crystal violet) for selective isolation of beta-haemolytic streptococci [21].

After 20–24 h incubation, presumptive beta-haemolytic colonies were streaked onto blood agar to obtain a pure growth. A catalase test was performed to differentiate the presumptive streptococci from staphylococci. Beta-haemolytic and catalase**-**negative isolates were grouped using the Streptex grouping kit (Remel R30950501) and ultimately reported as group A, B, C, D, F or G [21].

#### Antimicrobial Susceptibility Testing

Pure morphologically similar colonies were made into a suspension equal to 0.5% MacFarland’s standard and streaked evenly over the surface of Muller Hinton Agar (MHA). Antimicrobial resistance was evaluated using the disk diffusion method (15 ug azithromycin disk) and all resistant isolates(zones of inhibition ≤13mm) were confirmed using E-test (AZ 256,range 0.016-256ug/ml, Biomerieux) [21]. The CLSI 2016 guidelines were used to interpret azithromycin susceptibility results.

#### Whole genome sequencing

Extracted DNA from all GAS isolates (isolated at any timepoint) was used for WGS by Illumina Miseq. Paired end reads quality checked (FastQC) and trimmed for adaptor contaminant and low Q-score bases (Trimmomatic), followed by *de novo* assembly using k-mer settings of 21, 22, 55 and 77 (SPAdes) [24, 25]. Assembled genomes were checked for post-assembly quality (QUAST) [26]. Criteria for inclusion in further analysis were <500 contigs, 1.6-2.0Mb (*S. pyogenes*) or 2.0-2.4Mb (*Streptococcus dysgalactiae* subspecies *equisimilis*; SDSE) assembly length, and >90% reference genome coverage (*S. pyogenes*). Assembled genomes were annotated using Prokka and the core genome determined using Roary [27,28]. Antimicrobial resistance genes were identified using ABRicate with the Resfinder database [29.30]. *Emm*-typing was performed using the bioinformatics method described at https://github.com/BenJamesMetcalf and the CDC *emm*-typing database (https://www2.cdc.gov/vaccines/biotech/strepblast.asp). Core genome alignments were used to draw maximum likelihood phylogenetic trees (RAxML) with 1000 bootstraps [31]. For comparative phylogenetic analysis of SDSE isolates, complete genomes were obtained from NCBI Genome resource (https://ncbi.nlm.nih.gov/genome/genomes/823?) and from a previous study reporting SDSE expressing Lancefield group A (SDSE(A)) [32]. Phylogenetic trees were visualized using iTOL (https://itol.embl.de/). All sequence data are publicly available at the European Nucleotide Archive (project accession PRJEB36490).

### Data Management and Statistical Analysis

The data was double entered into Open Clinica and analysed using STATA 16. We only included participants with all samples collected at each of the specified timepoints. The prevalence of GAS carriage was determined as the presence at any time point for each of the sample types. Prevalence was compared between arms using prevalence ratios (PR) with their corresponding 95% confidence intervals (CI). We used Fisher exact test to obtain p-values. Prevalence of azithromycin resistance was calculated considering all sampled individuals as the denominator. If any isolate from a biological site was resistant, the sample of that participant was considered as resistant.

### Ethical Approval

Ethical approval for the main trial was obtained from the Joint MRC Gambia Government Ethics Committee. Ethical approval was given by LSHTM for the secondary data analysis.

## RESULTS

### Study Population

We recruited 829 women for the trial (414 women to the azithromycin and 415 to the placebo arms) who delivered a total of 843 babies, including 13 stillbirths. Overall, 715 pairs (86.2%) had all study samples collected and are part of this posthoc analysis (Figure 1). Most women were 20-29 years old and the major ethnic group was Mandinka. Approximately 66.6% of deliveries occurred during the dry season (November to May), 5.0% of the newborns were low-birth weight and slightly more than half were males (Table 1).

**Table 1:**
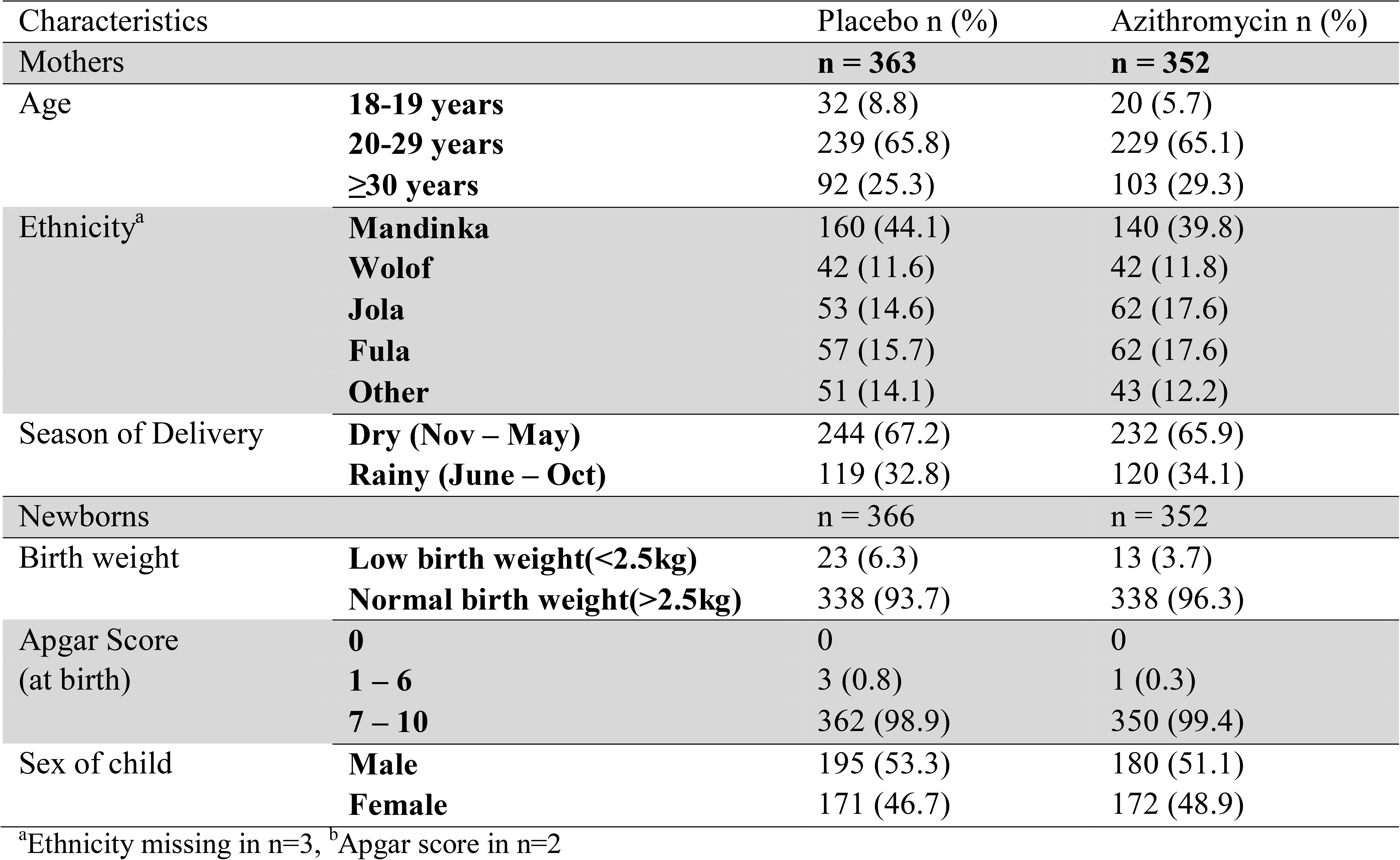
Baseline Demographic Characteristics of study participants

**Figure 1:**
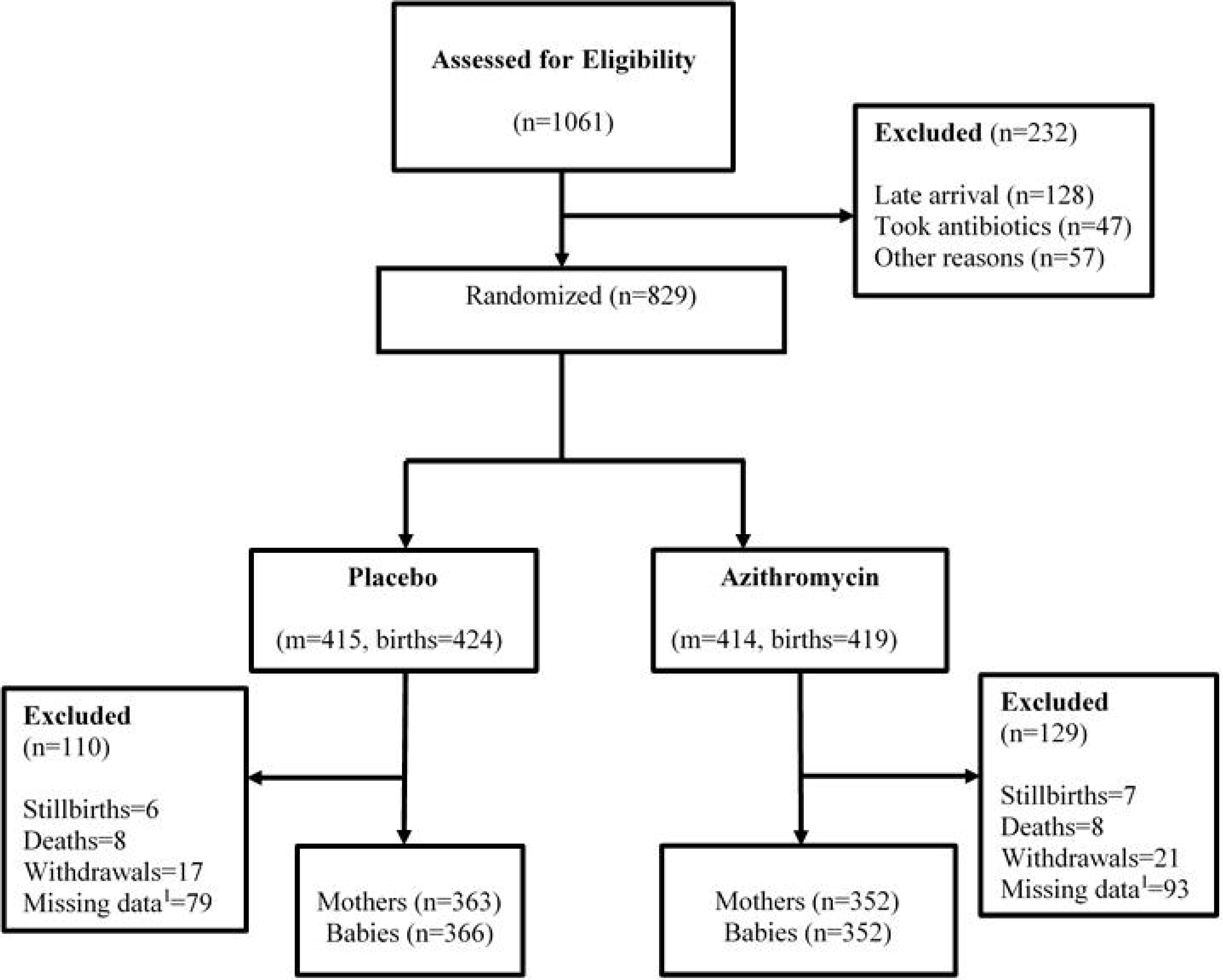
Trial profile ^1^ Mother/baby pair with ≥1 missing sample

### Prevalence of GAS carriage

Overall, 30 women and 9 newborns had at least 1 sample positive for GAS; 7 women and one baby had 2 positive samples and one woman had 5 positive samples (Total of 51 GAS isolates).

#### Study mothers

Pre-intervention prevalence of GAS was low and similar in the two study arms, both in the nasopharynx and in the vaginal tract (Table 2, Figure 2a). Post intervention, prevalence of GAS in the azithromycin arm was lower in the nasopharynx (0.28% versus 1.93%, PR=0.15 95% CI (0.02,1.19), p=0.069) and breast milk (0.28% versus 2.48%, PR=0.11 95% CI (0.01,0.90), p=0.021) but not in the vaginal tract (1.99% versus 1.93%, PR=1.03 95%CI (0.37,2.91), p=1.000) (Table 2).

**Table 2:** Prevalence of GAS carriage in the nasopharynx, breastmilk and vaginal samples of mothers and nasopharynx of babies

| **MOTHERS** |  |  |  |  |
| --- | --- | --- | --- | --- |
|  | Placebo (%)<br>n = 363 | Azithromycin (%)<br>n = 352 | Prevalence Ratio<br>(95% CI) | p-value |
| **Nasopharyngeal carriage** |  |  |  |  |
| Pre-intervention<sup>a</sup> | 0 | 1 (0.28) | - | 0.492 |
| Post Intervention<sup>b</sup> | 7 (1.93) | 1 (0.28) | 0.15 (0.02,1.19) | 0.069 |
| **Breastmilk carriage<sup>c</sup>** |  |  |  |  |
| Post Intervention<sup>b</sup> | 9 (2.48) | 1 (0.28) | 0.11 (0.01,0.90) | 0.021 |
| **Vaginal carriage** |  |  |  |  |
| Pre-intervention<sup>a</sup> | 1 (0.28) | 2 (0.57) | 2.06 (0.18,22.64) | 0.619 |
| Post Intervention<sup>d</sup> | 7 (1.93) | 7 (1.99) | 1.03 (0.37, 2.91) | 1.000 |
| **NEWBORNS** |  |  |  |  |
|  | Placebo (%)<br>n = 366 | Azithromycin (%)<br>n = 352 | Prevalence Ratio<br>(95% CI) | p value |
| Post intervention<sup>e</sup> | 7 (1.91) | 2 (0.57) | 0.30 (0.06,1.42) | 0.178 |

<sup>a</sup>Carriage at day 0 before treatment
<sup>b</sup>Carriage at days 3, 6, 14 and 28 combined
<sup>c</sup>There are no pre-intervention samples for the breast milk
<sup>d</sup>Day 10(day 8 – 13)
<sup>c</sup>Day 0, 3,6,14 and 28 combined. Day 0 is the day of birth

**Figure 2:**
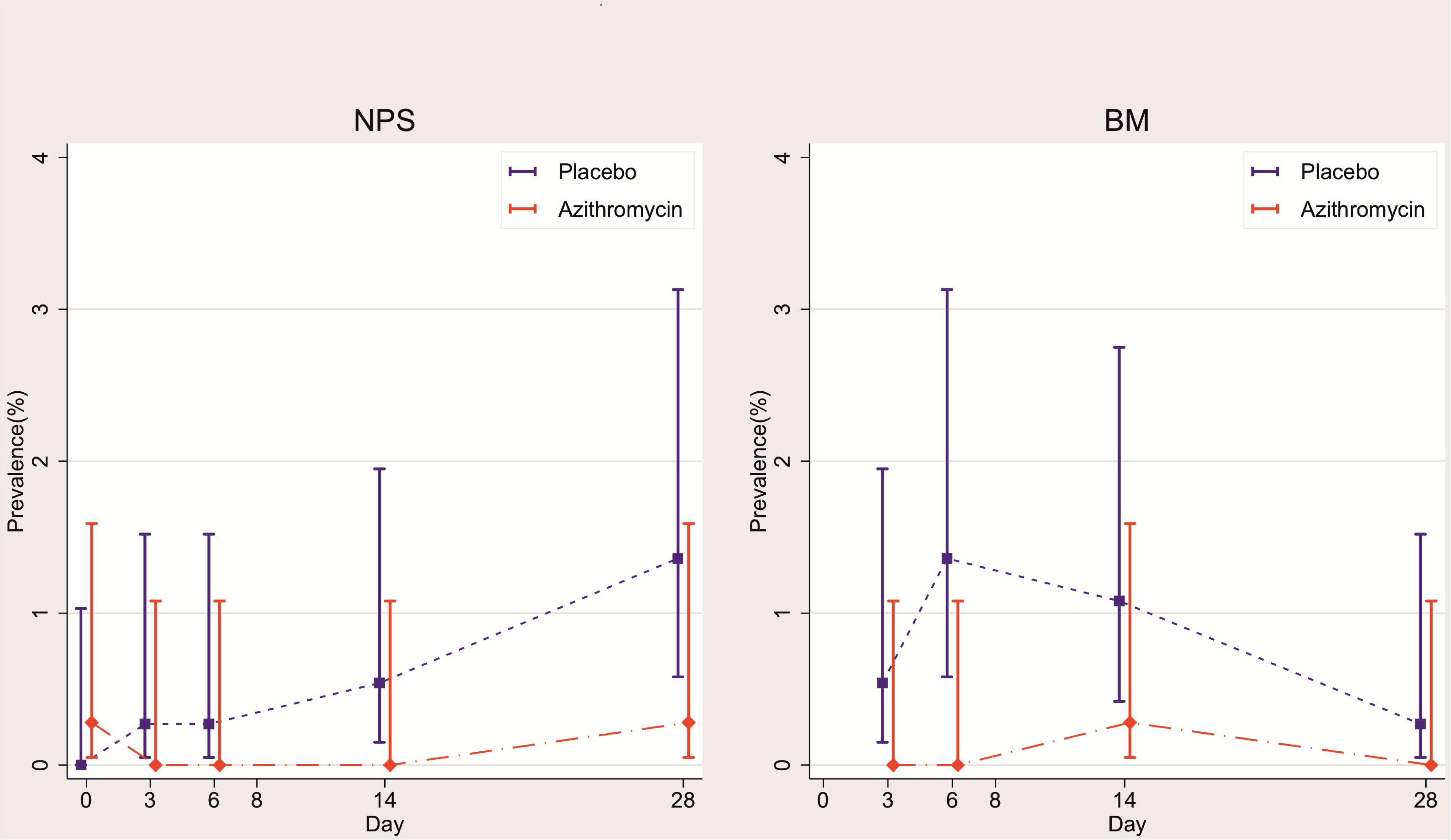
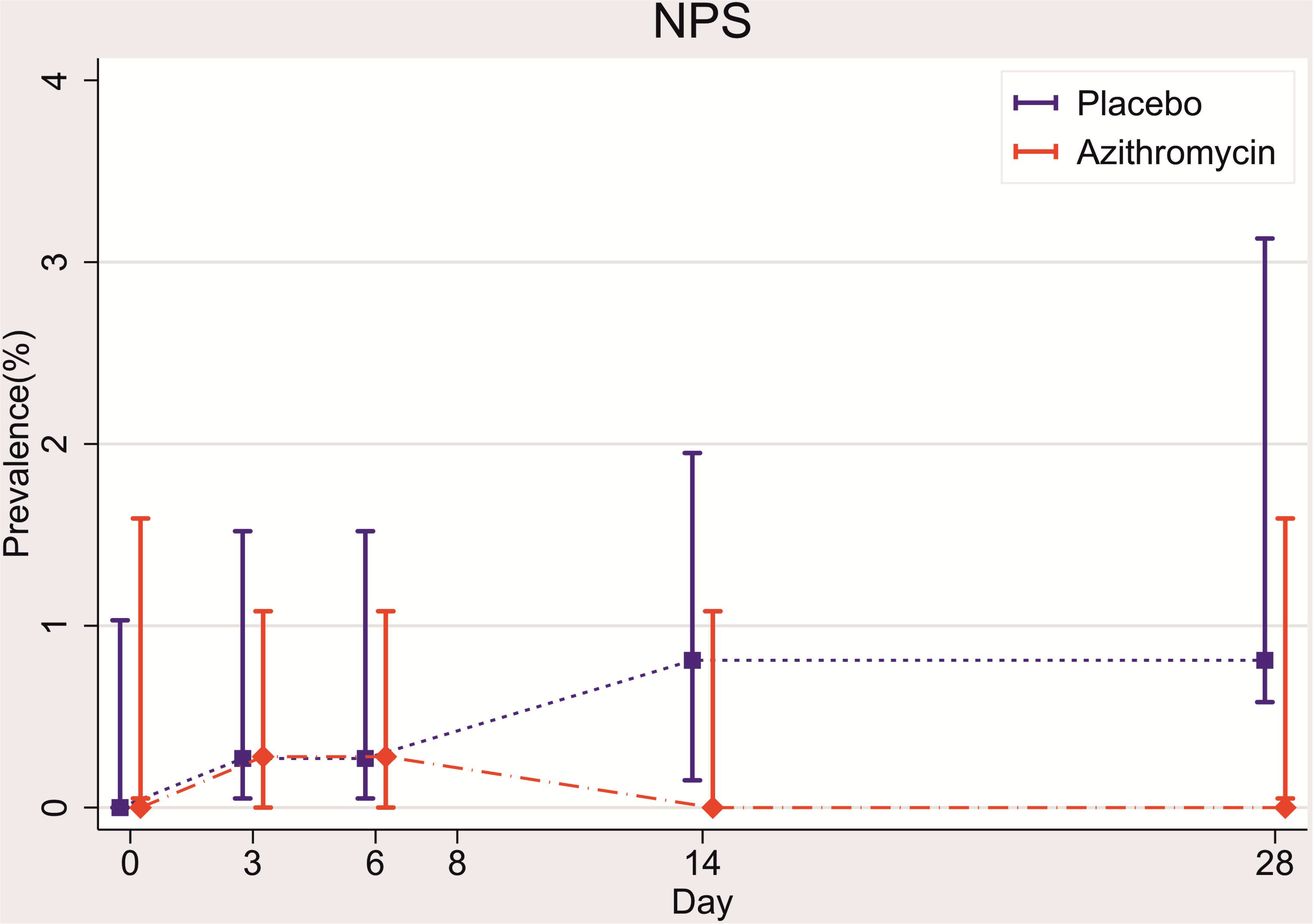
(a) Prevalence of i) maternal nasopharyngeal carriage and ii) breastmilk carriage of GAS at different timepoints in the azithromycin and placebo arms. (b) Prevalence of neonatal nasopharyngeal carriage of GAS at different timepoints in the azithromycin and placebo arms

#### Study newborns

Prevalence of nasopharyngeal carriage of GAS was low; with a lower prevalence in the azithromycin arm (0.57% versus 1.91%, PR=0.30 95%CI (0.06,1.42), p=0.178) (Table 2, Figure 2b).

### Prevalence of azithromycin resistance

#### Study mothers

Prevalence of azithromycin-resistant GAS was similar between study arms for all sample types except for higher resistance in vaginal GAS isolates in the intervention arm [1.99% vs. 0.28%, PR=7.24, 95% CI (0.87,56.92 p=0.035)] (Table 3).

**Table 3:** Prevalence of carriage of resistant GAS in the nasopharynx, breastmilk and vaginal samples of mothers and nasopharynx of babies. ^a^Carriage at day 0 before treatment

| **MOTHERS** |  |  |  |  |
| --- | --- | --- | --- | --- |
|  | Placebo (%)<br>n = 363 | Azithromycin (%)<br>n = 352 | Prevalence Ratio<br>(95% CI) | p-value |
| **Nasopharyngeal carriage** |  |  |  |  |
| **Pre-intervention<sup>a</sup>** | 0 | 0 | - | - |
| **Post-Intervention<sup>b</sup>** | 2 (0.55) | 1 (0.28) | 0.52 (0.05,5.66) | 1.000 |
| **Breastmilk carriage<sup>c</sup>** |  |  |  |  |
| **Post-Intervention<sup>b</sup>** | 0 | 1 (0.28) | - | 0.492 |
| **Vaginal carriage** |  |  |  |  |
| **Pre-intervention<sup>a</sup>** | 0 | 1 (0.28) | - | - |
| **Post-Intervention<sup>d</sup>** | 1 (0.28) | 7 (1.99) | 7.24 (0.87,56.92) | 0.035 |
| **NEWBORNS** |  |  |  |  |
|  | Placebo (%)<br>n = 366 | Azithromycin (%)<br>n = 352 | Prevalence Ratio<br>(95% CI) | p value |
| **Post-intervention<sup>e</sup>** | 1 (0.27) | 1 (0.28) | 1.04 (0.07,16.56) | 1.000 |

<sup>b</sup>Carriage at days 3, 6, 14 and 28 combined
<sup>c</sup>There are no pre-intervention samples for the breast milk
<sup>d</sup>Day 10(day 8 – 13)
<sup>e</sup>Day 0, 3,6,14 and 28 combined. Day 0 is the day of birth

#### Study newborns

Prevalence of GAS azithromycin resistance was low and similar between arms with only two participants having resistant GAS isolates (Table 3).

### Whole Genome Sequencing

Of the 51 GAS isolates, one sample was not retrieved and five samples (isolated from four mothers) had poor sequences and were excluded from the WGS analysis. Of the remaining 45 isolates, WGS confirmed that 35 were *S. pyogenes* (2 from the azithromycin arm and 33 from the placebo arm of the trial), from which we detected 16 *emm* types. The most common were *emm*4 and *emm*44 (5 isolates each, 14.2%) and *emm*147 (4 isolates, 11.4%). The remaining 10 isolates were SDSE. All were phenotypically resistant to azithromycin and 9 were from participants in the azithromycin arm. All 10 SDSE(A) isolates were retested with Streptex grouping kit and were confirmed as Lancefield group A beta-haemolytic streptococci. We observed that the most common resistance mechanism was by efflux with 14 out of 16 azithromycin-resistant isolates (including all SDSE(A)) harbouring both mefA and msrD genes (mefA-msrD). In *S. pyogenes*, mefA and msrD genes were adjacent to each other and located five genes upstream of catQ on what appeared to be a phage-like mobile genetic element, integrated downstream of rlmD (23s rRNA methyltransferase). In SDSE(A), mefA and msrD were also present on a mobile genetic element that showed some similarity to that found in the reference SDSE strain AC-2713 (HE858529.1) integrated between comEC and comEA, but differed in gene content between the two lineages of SDSE(A). The complete sequences of the mobile elements for both *S. pyogenes* and SDSE(A) could not be determined due to contig breaks in the *de novo* assemblies. *S. pyogenes* isolates recovered from the same mother from different biological sites or study timepoints, as well as isolates from their new-borns, all clustered together and belonged to the same *emm* type (Figure 3a). Similar phylogenetic and epidemiological concordance was seen in the SDSE(A) isolates, including a neonatal NPS isolate closely linked to those recovered from the baby’s mother (Figure 3b).

**Figure 3:**
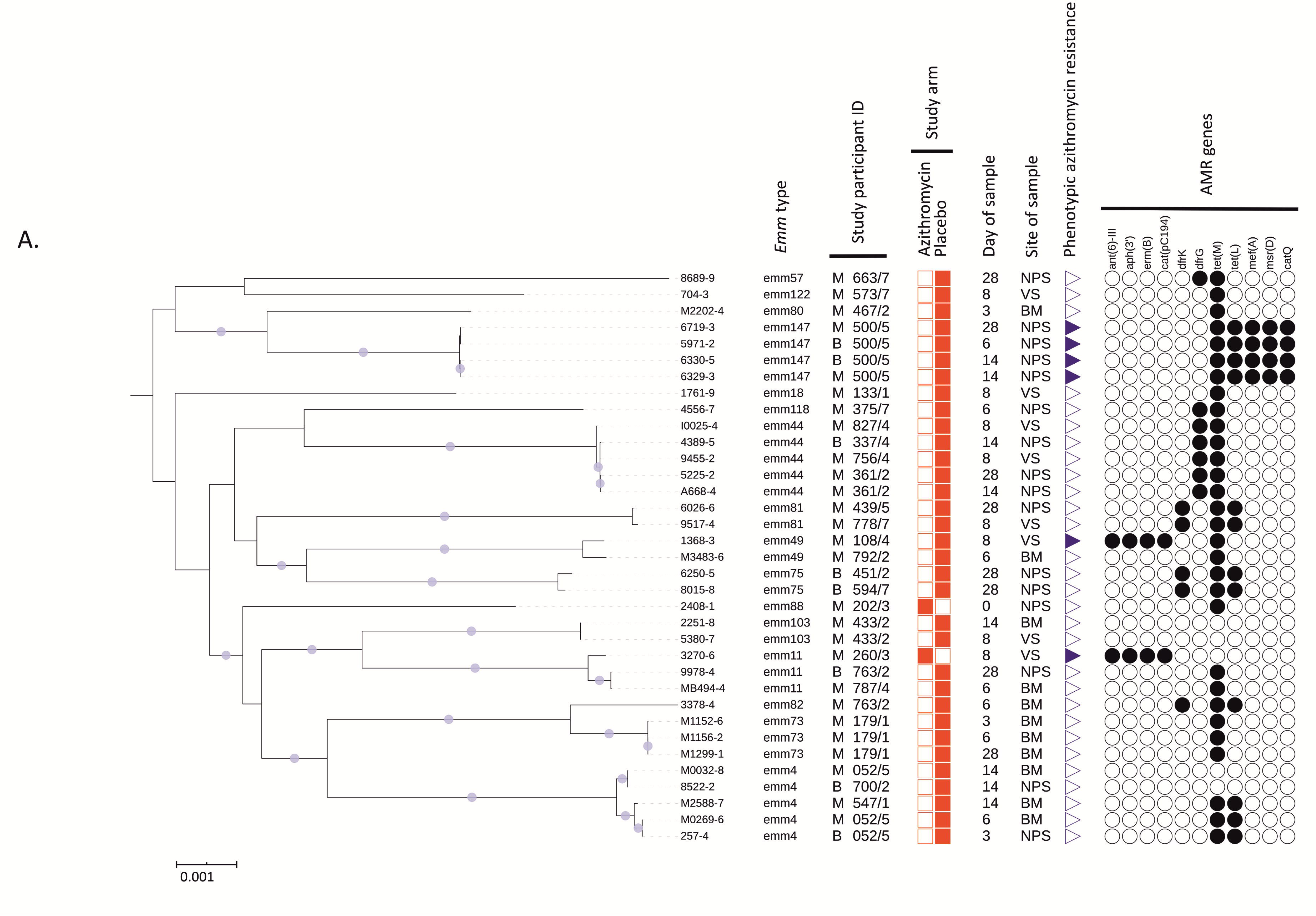
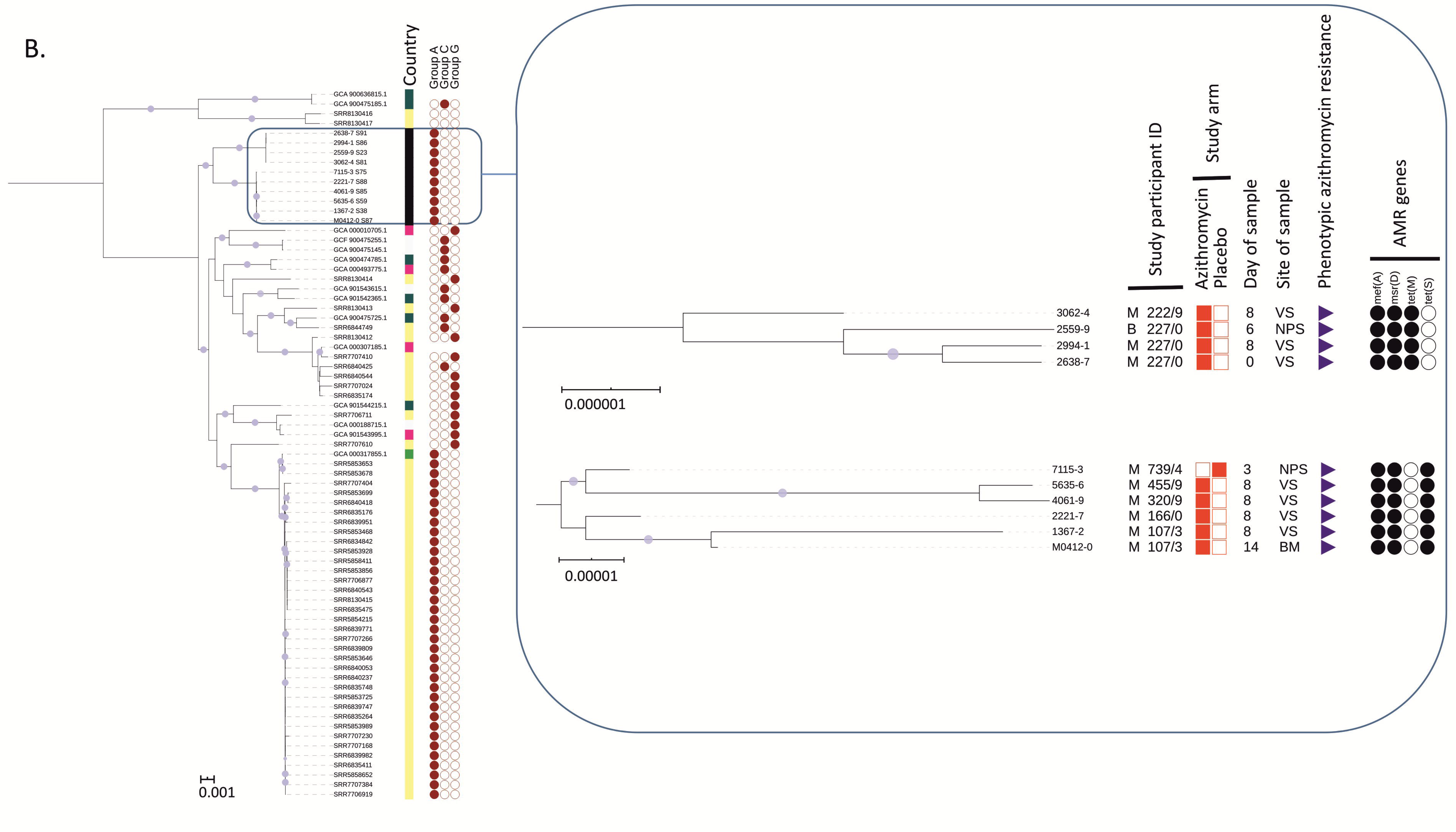
Midpoint rooted maximum likelihood core-genome phylogenetic analysis using RAxML GTRCAT model with 1000 bootstrap replicates. Circle symbols indicate >99% bootstrap support. (a) Core-genome (1299 genes) phylogenetic analysis of 35 *S. pyogenes* isolates from the study cohort. (b) International contextualization (based on core genome of 1221 genes) of 10 *S. dysgalactiae* subspecies *equisimilis* isolates with individual core-genome (upper clade: 2106 genes, lower clade: 2078 genes) phylogenetic analysis of the two distinct clades in the study cohort. (Annotation key: country of origin; black=the Gambia, yellow=USA, dark green=UK, light green=Germany, pink=Japan, white=unknown, symbols; filled=present, unfilled=absent, no symbol=unknown; study participant ID (unique study identifier for mother/baby units); M=mother, B=newborn, NPS=nasopharyngeal swab, VS=vaginal swab, BM=breast milk, AMR=antimicrobial resistance).

## DISCUSSION

One oral dose (2g) of azithromycin given to women in labour reduced the prevalence of GAS carriage among women and their babies in the nasopharynx and breast milk without an increase of azithromycin resistance in isolates in these sample sites. In contrast, the intervention did not have any effect on the prevalence of GAS carriage in the vaginal tract but induced an increase in the carriage prevalence of azithromycin-resistant SDSE(A).

We have previously shown that a single oral dose (2g) of azithromycin given to women in labour reduced the prevalence of *S. aureus, S. pneumoniae* and GBS carriage in the mother (nasopharynx, breast milk and vaginal tract) and the baby (nasopharynx) [22]. The current analysis shows that the intervention also reduced the prevalence of GAS carriage in the breast milk and nasopharynx of study women and, less clearly, in the nasopharynx of their newborns. Despite substantial reduction of GAS carriage, we did not observe any short-term increase of azithromycin resistance in these two biological sites.

Conversely, the intervention did not have any effect on the prevalence of GAS carriage in the maternal vaginal tract but induced an increase in azithromycin resistance. WGS revealed that GAS vaginal carriage in the azithromycin arm was primarily due to azithromycin-resistant SDSE(A), whereas in the placebo arm, GAS vaginal carriage was entirely due to azithromycin-susceptible *S. pyogenes*. In our previous analysis on *S. aureus*, lower reduction in carriage in the vaginal tract alongside a higher prevalence of resistance was also observed [22]. It is not clear why the effect of the intervention in the vaginal tract differs from other body sites. The concentration of azithromycin in the vaginal tract may be lower and fall more rapidly than in other biological sites. We had previously shown a very high concentration of azithromycin in breast milk during the 4 weeks following the intervention, with a peak during the first 6 days (concentration >4,000 µg/L) [33]. A different study using a single dose of azithromycin (1g) showed the azithromycin concentration in the vaginal tract was much lower than the breast milk concentration we observed [34]. In this study, the peak concentration occurred during the first 24-48 hours following the intervention [34], long before the post-intervention VS were collected in our study. It is possible that removal of *S. pyogenes* from the vaginal tract allows azithromycin-resistant SDSE to thrive, whereas in other body sites higher concentrations of azithromycin can overcome the efflux-mediated resistance mechanisms [35]. An alternative explanation is that even though SDSE can be found in different biological sites, it is better suited to survive in the vaginal tract [36]. One azithromycin-resistant SDSE(A) was isolated in the vaginal tract from a woman included in the azithromycin arm before the intervention was administered. A phylogenetically-linked isolate was isolated from the same woman’s VS after the intervention.

The public health and clinical implications of the selective expansion of SDSE(A) in the vaginal tract are difficult to anticipate. However, similarly to *S. pyogenes*, SDSE can cause invasive disease [37-39]. Lancefield group A SDSE has been described in previous studies from high income settings, including a collection of isolates causing invasive disease in the USA [32]. The SDSE(A) isolated from our study participants have distinct phylogeny to SDSE(A) previously isolated in USA and fall into two distinct clades. In our study, all SDSE(A) isolates harboured *mefA-msrD* genes, whereas both *erm(B)* and *mefA-msrD* genes were found in azithromycin-resistant *S. pyogenes* isolates. While the presence of *mefA* is associated with macrolide resistance, *msrD* has a more dominant role [40, 41]. The presence of both *mefA* and *msrD* may confer high level resistance [41].

The trial was designed to evaluate the effect of intra-partum azithromycin on maternal and neonatal carriage of *S. aureus, S. pneumoniae* and GBS that are more prevalent than *S. pyogenes* and therefore the current analysis was underpowered as observed in the comparison of trial arms in the nasopharyngeal swabs, especially in newborns. This was an opportunistic study and oropharyngeal rather than nasopharyngeal samples may have been more appropriate for detecting *S. pyogenes* carriage [42], but adds to the growing evidence that group A *Streptococcus* may include SDSE as well as *S. pyogenes*.

In conclusion, this study demonstrates that a simple intervention (single dose of intra-partum oral azithromycin), besides reducing carriage of *S. aureus, S. pneumoniae* and GBS, can also reduce carriage of GAS, an important cause of maternal and neonatal sepsis. We need to closely monitor the effect of this prophylactic intervention on azithromycin-resistant SDSE(A) isolates and its effect on disease when assessing the overall public health potential of prophylactic intra-partum azithromycin.

## Supporting information

Consort checklist

## Data Availability

Access to data should be approved by the internal MRCG at the LSHTM committee (Scientific Coordinating Committee) which meets on monthly bases. Requests should be submitted to the Government department.

## Supplementary Data

Supplementary 1: WGS_data.xlsx

## Author’s contributions

AR and UDA conceived and designed the initial trial. AR, IJ and TdS conceived this posthoc study. IJ wrote the manuscript and AR, TdS, AJK contributed significantly in the final version. CO, BC, AB developed and adapted the field and laboratory work. IJ, EJ and ES analysed the samples. AKS led the team that conducted the WGS. AJK, SYB, CET and TdS performed the WGS analysis. CB provided statistical input to the study. All authors read and approved the final manuscript.

## Acknowledgements

The authors express our profound gratitude to the study participants and their families for agreeing to take part in the study. We give special thanks to the laboratory team (Njemmeh Manneh, Aji Mary Taal, Aru Kumba Baldeh, Modou Lamin Fofana, Momodou Abass Bah and Nano Kora) for their involvement in sample reception and processing. We also extend our gratitude to Jarra Manneh and Abdoulie Kanteh of the MRCG genomics team, Nuredin Ibrahim Mohammed for assisting with generating some figures and the field team (led by Edrissa Sabally and Omar Jarra) for assisting with sample collection.

